# Role of interfering substances in the survival of coronaviruses on surfaces and their impact on the efficiency of hand and surface disinfection

**DOI:** 10.1101/2020.08.22.20180042

**Authors:** L Szpiro, A Pizzorno, L Durimel, T Julien, A Traversier, D Bouchami, Y Marie, M Rosa-Calatrava, O Terrier, V Moules

## Abstract

Contaminated environmental surfaces are considered to represent a significant vector for hospital-acquired viral infections. In this study, we have evaluated the impact of interfering substances on SARS-CoV-2 surface stability and virucidal efficiency of hand sanitizers and surface disinfectant. To this end, surface stability of SARS-CoV-2 was measured on stainless steel in different experimental conditions, with or without an artificial mucus/saliva mixture and compared against that of human coronavirus HCoV-229E and feline coronavirus FCoV. The impact of the mucus/saliva mixture on the virucidal efficiency of 3 commercial alcohol hand sanitizers and 1 surface chemical disinfectant against SARS-CoV-2, HCoV-229E and FCoV was then measured. Our results indicate that mucus/saliva mixture did not demonstrate a beneficial effect on the surface survival of tested viruses, with temperature being an important parameter. In addition, we demonstrated that interfering substances may play an important role in the virucidal efficacy of hand sanitizers and disinfectants, highlighting the need for adapted testing protocols that better reflect current “real life” conditions of use.

**Highlights:**

- Contaminated environmental surfaces are a significant vector for viral infections.
- We studied the impact of interfering substances on SARS-CoV-2 surface stability and virucidal efficiency.
- Mucus/saliva did not demonstrate a beneficial effect on viral surface stability, with temperature being an important parameter.
- Interfering substances are important for virucidal surface activity of disinfectants.

## Introduction

Respiratory diseases caused by human coronavirus infection are of both medical and socio-economic importance. Severe acute respiratory syndrome coronavirus (SARS-CoV), Middle East respiratory syndrome coronavirus (MERS-CoV) and currently SARS-CoV-2 have crossed the species barrier and entered the human population to cause severe disease. As observed for a large number of respiratory viruses, airborne transmission and fomite transmission are thought to play important roles in the epidemiology of these viruses (1). Several reviews and models have suggested that indirect contact transmission involving contaminated surfaces could be the predominant transmission route for certain respiratory viruses, including influenza, in some settings (2–4). Indeed, contaminated environmental surfaces are considered to represent a significant vector for hospital-acquired viral infections (5, 6). Surface contamination by respiratory viruses may occur by direct landing of droplets expelled during the coughing or sneezing of an infected individual or as a result of indirect transfer from their hands contaminated with excreted virus. During the 2002-2003 SARS epidemic, the detection of virus on a variety of surfaces and objects in healthcare settings provided evidence of fomites playing an important role in viral dissemination (7). Boone and Gerba (2) demonstrated the presence of influenza virus on 50% of fomites found in homes and daycare centers during an active influenza season. Influenza A virus has also been shown to be transferred from stainless steel countertops to hands up to 24 h after surface contamination (8). The transfer of human parainfluenza virus and rhinovirus from contaminated surface to clean fingers supports a role for fomites in the contamination of hands with both viruses (9).

For any environmental contamination to be relevant, a virus should not only remain infectious on the recipient surface but also persist at a sufficient concentration to enable it to reach the respiratory tract via finger contamination. In general terms, the potential of a fomite to spread a given infectious agent is directly related to the capacity of the agent to survive on that surface. The surface stability of viruses is generally influenced by the type of surface, environmental factors such as relative humidity (RH) and temperature, and the presence of body fluid secretions.

Coronaviruses have the capacity to survive on a wide range of porous and non-porous materials (10). A recent study has shown that SARS-CoV and SARS-CoV-2, at 40% RH and 21-23 °C, were most stable on smooth surfaces and that viable virus could be detected up to 3 and 4 days respectively post application (10). Chin et al showed that SARS-CoV-2 could be detected on contaminated stainless steel supports, at 65% RH and 22 °C, seven days after experimental inoculation (11). SARS-CoV has been reported to stay viable for up to five days at 22 to 25 °C and 40 to 50% RH, with an increase in temperature and humidity resulting in a rapid loss of viability (10). Casanova et al demonstrated a higher stability in SARS-CoV compared to the human coronavirus HCoV-229E, which in a dried state retained residual infectivity even after 6 days compared to the loss of infectivity within 3 days observed in HCoV-229E (12). MERS-CoV virus could still be recovered on stainless steel after 2 days in the 20 °C and 40% RH condition, whereas it remained viable for a much shorter 8 h at 30 °C and 80% RH, and 24 hours at 30 °C and 30% RH (13). All studies that have tested varying temperature and RH agree that lower temperatures and higher RHs systematically favor the survival of coronaviruses (13,14).

During and after illness, viruses are shed in large numbers in body secretions, including blood, feces, urine, saliva, and nasal fluid, all of which comprise high levels of protein and other biological organic matter. While several studies appear to have reached a consensus agreement on the role of both fecal material and blood in the increased stability of viruses on environmental surface (15–17), the potential beneficial effect of respiratory secretions on surface viral survival remains unclear. Mixing of highly concentrated inocula with respiratory mucus increased the infectiousness of influenza viruses, allowing its transmission for up to 17 days (18) and thus confirming the protective role of human mucus for the survival of respiratory viruses previously shown by Parker et al (19). However, human rhinovirus type 14 suspended in tryptose phosphate broth could survive for more than 20 h incubation, though for less time when suspended in bovine mucin or in nasal secretions (20).

No data are available concerning the transmissibility of coronaviruses from contaminated surfaces to hands. Data are however available concerning other respiratory viruses including influenza A virus, parainfluenza virus 3 and rhinovirus which were shown to be transferred to the hands (1.5 to 31.6% of the viral load) after a contact of 5 s with an inoculated surface (24, 9). Several studies have found that frequent hand-surface contact can lead to rapid diffusion of surface contamination (25, 26). In view of the indirect transmissibility of viruses, compliance with both hand hygiene and surface disinfection recommendations is critical to reducing colonization and infection of the hands of entire populations but in particular the hands of health-care workers.

An important consideration in the strict compliance of these recommendations is the finding in several studies that interfering substances (including body secretions) could have a strong impact on the efficiency of a disinfectant (27, 28). To simulate clean/dirty environments, standardized interfering substances are used to evaluate disinfection product efficiency according to Phase 2 EN standards. While the determination of surface virucidal efficiency of a disinfectant is managed according to EN16777 standards through the use of inoculated stainless steel carriers, the virucidal efficiency of hand rub products is currently tested using the EN14476 suspension test (29, 30). The efficiency of hand rub products is based upon a titer reduction of ≥ 4 log10 in a maximum contact time of 30 s to 60 s in compliance with suspension test standards.

In an attempt to better understand and thus better control the transmission of SARS-CoV-2 behind the recent and ongoing pandemic, the impact of body fluid secretions, from coughing or sneezing corresponding to an artificial mixture containing nasal mucus and saliva, on the surface stability of SARS-CoV-2 and the virucidal efficiency of disinfectant were tested. To this end, surface stability was measured on stainless steel at 7 and 25 °C (65% RH) with or without the artificial mucus/saliva mixture and compared against that of human coronavirus HCoV-229E and the standardized feline coronavirus FCoV strain used in EN standard tests of surface virucidal activity (23). The impact of the mucus/saliva mixture on the virucidal efficiency of 3 commercial alcohol hand sanitizers (according to the EN14476 standard suspension test) and 1 surface chemical disinfectant (according to the virucidal surface quantitative EN16777 test) against SARS-CoV-2, HCoV-229E and FCoV was then measured.

## Methods

### Viruses and cell lines

The HCoV-229E coronavirus (ATCC-VR-740), the SARS-CoV-2 (BetaCoV/France/IDF0571/2020) and the feline coronavirus (FCoV, RVB-1259) were chosen for this study. HCoV-229E, SARS-CoV-2 and FCoV were produced on MRC5 cells (ATCC-CCL-171), on Vero E6 cells (ATCC- CCL-1586) and on CRFK cells (CCLV-RIE-115), respectively. For the evaluation of surface disinfectant, another enveloped virus was introduced, the modified vaccinia virus Ankara (MVA, ATCC VR-1566), produced on BHK-21 cells (ATCC CCL-10). Cells were cultivated in DMEM (1 or 4.5 g/l of glucose) or EMEM medium with L-glutamine and antibiotics, with or without FCS.

### Virus stability

The stability was evaluated on functionalized stainless steel disks. Disks were washed in 95% Decon then rinsed with distilled water and immersed in 70% ethanol before autoclaving 10 min at 121°C. Virus stability on stainless steel disks was determined by the recovery of virus previously deposited on surface. The inoculum was made by following the NF EN 16777 standard. 50 μL of purified virus or virus mix with artificial saliva (ASTM 2721, Table 1) and epithelium mucus was put in the center of a stainless-steel disk. The deposit was dried under PSM for a maximum of 1 h. Disks were then placed in a refrigerator at 7 °C or in an oven stabilized at 25 °C and 65% relative humidity for different lengths of time (from 0 to 96 h). The recovery of each virus at different time points was performed using the same protocol: 2 ml of infection medium was first placed on the disk and incubated for 3 min at room temperature. All the liquid was collected into a tube and quantified by TCID50/ml (Spearman and Karber method). The quantification limit was determined taking into account the cytotoxicity. The limit of detection (LOD) on every cell, was 0.5 log10 TCID50/support.

### Virucidal activity according to suspension test

The virucidal activity of three commercial hand rub products against SARS-CoV-2, HCoV-229E and FCoV was determined using the quantitative suspension test according to EN 14476, comparing standardized interfering substance (clean condition, 0.3 g/l BSA) and our artificial mucus/saliva mixture. A 0.1 part by volume of virus suspension and 0.2 parts by volume of interfering substance were mixed with 9.7 parts by volume of the hand rub product corresponding to the modified protocol to evaluate ready-to-use products at 97% according to the EN14476 standard. At the tested contact time, a tenfold dilution was made to stop the activity of the disinfectant before quantifying the remaining infectious virus using the Spearman and Karber method. Contact times of 30 and 60 s were assessed at 20 °C. Titer reduction is presented as the difference between the control virus titer and that after contact with the test product. This difference is given as a reduction factor including its 95 % confidence interval. A reduction in virus titer of ≥ 4 log10 (corresponding to an inactivation of ≥99.99 %) was regarded as evidence of sufficient virucidal activity according to EN14476 standard. The quantification limit was determined taking into account the cytotoxicity (2.5 log10 TCID50). All tests were performed in duplicate and included all EN14476 controls.

### Virucidal activity according to surface test

To measure the virucidal activity on surface of a commercial disinfectant, the NF EN 16777 standard protocol was followed. The purified virus was mixed with the interfering substance (BSA 0.3 g/l, artificial saliva + epithelium mucus, BSA 3g/l + 3 ml/l of sheep erythrocytes). A volume of 50 μl of the mix was deposited on a previously functionalized stainless-steel disk (see previously). The inoculum was left to dry before 0.1 ml of the disinfectant was deposited on the disk. The remaining infectious viruses were quantified by the Spearman and Karber method. The quantification limit was determined taking into account the cytotoxicity (1.5 log10 TCID50). All tests were performed in duplicate and included all EN16777 controls.

## Results

### Comparative surface stability of SARS-CoV-2, HCoV-229E and FCoV

To investigate the stability of SARS-CoV-2, HCoV-229E and FCoV on stainless steel at both 7 and 25 °C, a 50-μl droplet of solution containing virus culture diluted in either culture media or artificial mixture (6.2, 7.1 and 6.8 log10 TCID50/ml respectively for SARS-CoV-2, HCov-229E and FCoV) was pipetted onto stainless steel discs and left in an incubator at 7 or 25 °C (relative humidity: 65%). The inoculated supports retrieved at desired time points were immediately soaked with 2 mL of viral culture media.

On stainless steel, SARS-CoV-2 showed greater stability at 7 °C than at 25 °C, with a loss at 48 h post application of 1.5 log10 TCID50 at 7 °C compared to 2.9 log10 TCID50 at 25 °C (**Figure 1A**). At 72 h post application, a 3.0 Log10 TCID50 reduction was observed at 7 °C while no viable virus could be detected at 25 °C. No viable virus could be measured on the surface at 7 °C at 96 h post application (**Figure 1A**). Both HCoV-229E and FCoV showed greater stability over time on stainless steel surface than SARS-CoV-2, with 1.7 log10 TCID50 reduction measured 96 h post application at 7 °C (**Figures 1B** and **1C**). As observed for SARS-CoV-2, the viral stability decreased at 25 °C with only a few (HCov-229E) or no (FCoV) viable virus being detected 96 h post application.

**Figure 1.**
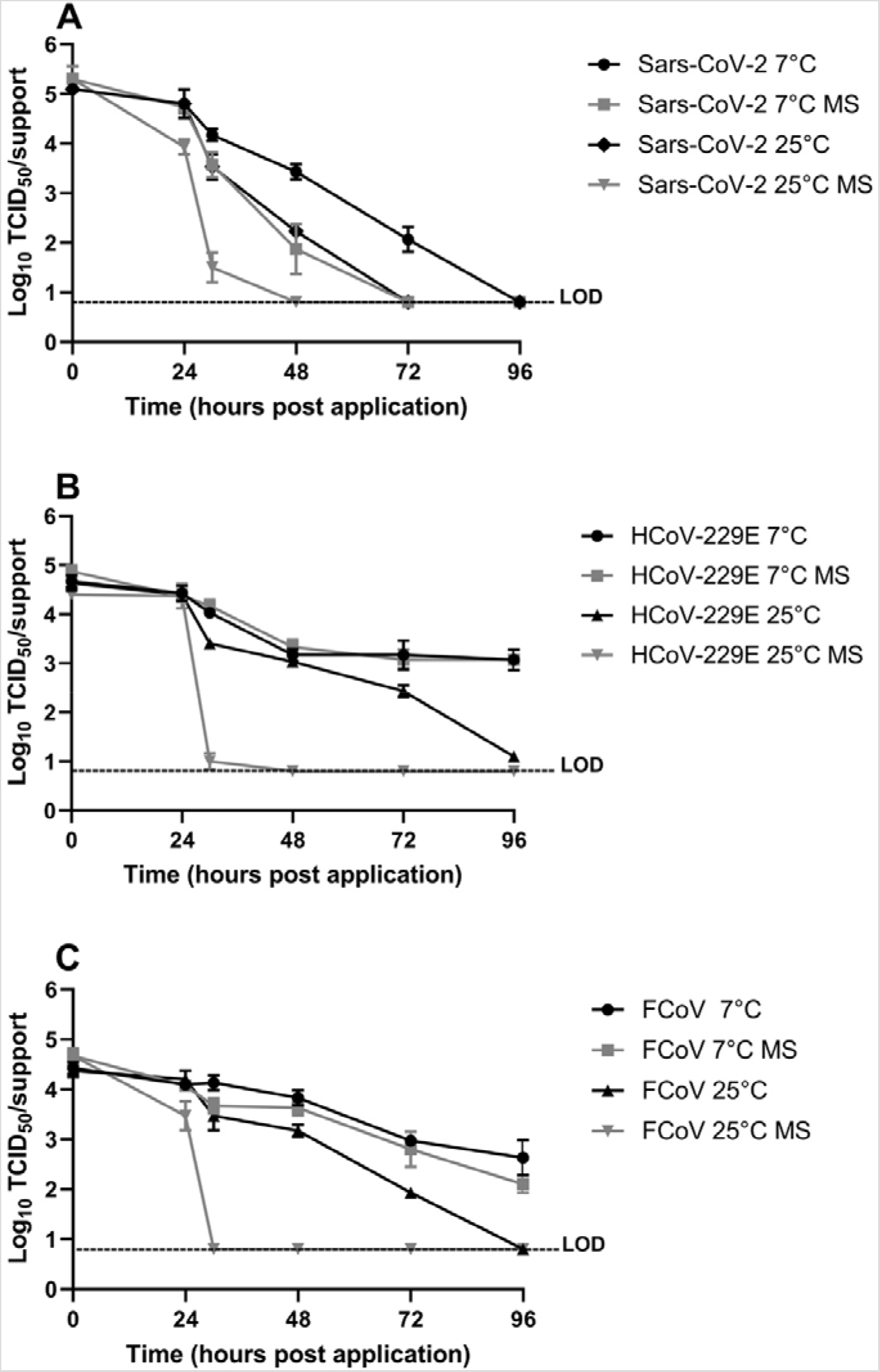
Stability of SARS-CoV-2 (A), HCov-229E (B) and FCoV (C) on stainless steel. Viral suspensions diluted in either culture media or artificial mucus/saliva mixture (6.2, 7.1 and 6.8 log10 TCID50/mL respectively for SARS-CoV-2, HCov-229E and FCoV) were deposited onto stainless steel discs and left in an incubator at 7 or 25 °C (relative humidity: 65%). The inoculated supports retrieved at desired time points were immediately soaked with 2 ml of viral culture media. The limit of detection (LOD) for all experiments was 0.8 Log10 TCID50/support.

### Effect of artificial mucus/saliva mixture on virus stability

We then evaluated the effect of an artificial mucus/saliva mixture on the stability of the three viruses under the same temperature conditions described above. The interfering mixture induced a moderate negative impact on the stability of SARS-CoV-2 at 7 °C, with 0.5, 1.6 and 1.9 log10 TCID50 reductions at 30, 48 and 72 h post application, respectively, compared to the condition without the artificial mixture (Figure 1A). This negative impact was even stronger at 25 °C, with no viable virus measured at 48 h post application. Artificial mucus/saliva mixture had no significant impact on the stability of HCoV-229E and FCoV at 7 °C (**Figures 1B** and **1C**), however, as observed for SARS-CoV-2, it had a strong negative impact in both at 25 °C, with no viable virus measured at 30 h post application (**Figures 1B** and **1C**).

### Virucidal activity according to suspension test

In order to measure the virucidal activity of alcohol hand sanitizers, the NF EN 14476+A2 standard protocol was followed. Briefly, a volume of 0.1 ml of interfering substance (BSA 0.3 g/l or artificial saliva/epithelium mucus) was added to 0.2 ml of purified virus and to 9.7 ml of disinfectant (ready-touse product). At the tested contact times (30 and 60 s), a tenfold dilution was made to stop the activity of the disinfectant before quantifying the remaining infectious virus.

Of three commercial hand rub products tested, only product 3 showed a virucidal activity at 30 s log10 against SARS-CoV-2 and all other tested viruses (**Figure 2A**), with the mucus/saliva mixture having no impact on virucidal efficiency (**Figure 2B**). Moreover, when compared to the standardized clean condition, the artificial mixture had no impact on the virucidal efficiency of either product 1 or 2. Product 1 demonstrated lower virucidal efficiency compared to product 2, with 2.3, 2.75 and 3.1 log10 reductions on SARS-CoV-2, HCoV-229E and FCoV titers, respectively, after a contact time of 30 s (**Figure 2A**). Of note, whereas a virucidal activity ≥4 log10 reduction was observed after a contact time of 60 s with products 1 and 2 against all three coronavirus strains, product 1 demonstrated a≥ 4 log10 reduction against SARS-CoV-2 in only one test after a contact time of 60 s (**Figure 2A**).

**Figure 2.**
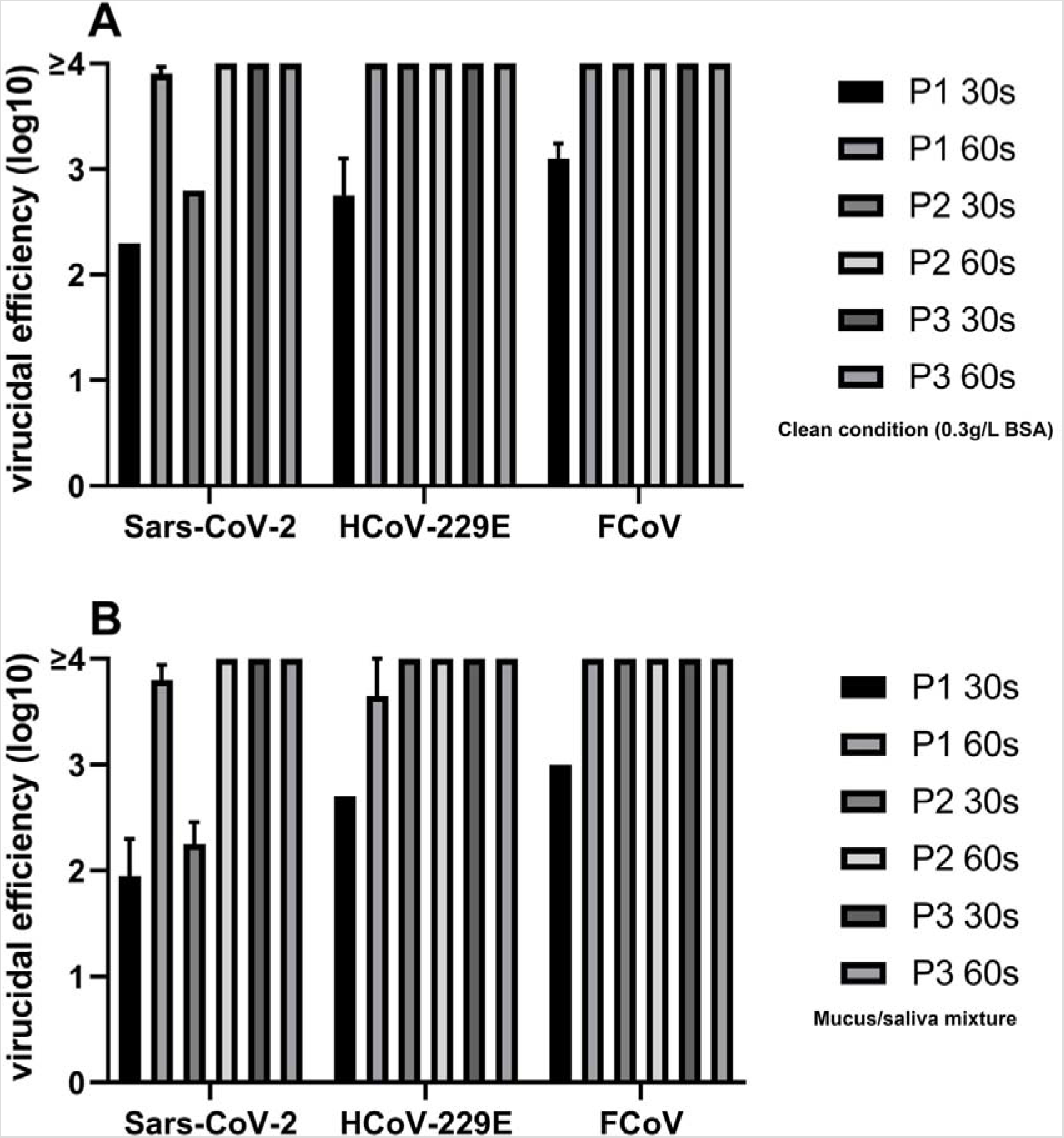
Virucidal activity of three commercial hand rub sanitizer products against SARS-CoV-2, HCoV-229E and FCoV. Virucidal activity was determined using the quantitative suspension test according to EN 14476, comparing the standardized interfering substance (“clean” condition, 0.3 g/l BSA (A) with the artificial mucus/saliva mixture (B). The remaining infectious viruses were quantified by the Spearman and Karber method. The quantification limit was determined taking into account the cytotoxicity of the products (2.5 log10 TCID50).

### Virucidal activity according to surface test

To further investigate a potential impact of the mucus/saliva mixture on virucidal efficiency, we evaluated the virucidal activity of a commercial chemical surface disinfectant using both clean/dirty medical standardized interfering substances (0.3 g/l BSA and 3 g/l BSA + 3 ml/l sheep erythrocytes) and the artificial mucus/saliva mixture following EN 16777 quantitative non-porous surface tests. Stainless steel discs were inoculated with 50 µl of a test suspension containing the standard modified vaccine Ankara virus strain (MVA) in a solution of interfering substance and let dry for 30 min. The dried inoculum was covered with 100 µl of surface disinfectant at 3 different concentrations (1, 2 and 4%) and maintained at 20 °C for a contact time of 2 min. A reduction in virus titer of ≥4 log10 was regarded as evidence of sufficient virucidal activity according to EN16777 standard.

All tested concentrations of the surface disinfectant showed virucidal activity ≥ 4 log10 after 2 minutes in contact with the MVA virus, with low levels of soil content corresponding to the standardized medical “clean” condition (**Figure 3**). However, the use of standardized medical “dirty” interfering substances (3 g/l BSA + 3 ml/l sheep erythrocytes) significantly decreased the virucidal efficiency of the product, with only the 4% concentration showing ≥ 4 log10 reduction. Interestingly, the mucus/salvia mixture had a similar impact on the virucidal efficiency on stainless steel surface than that observed with the standardized “dirty” condition (**Figure 3**).

**Figure 3.**
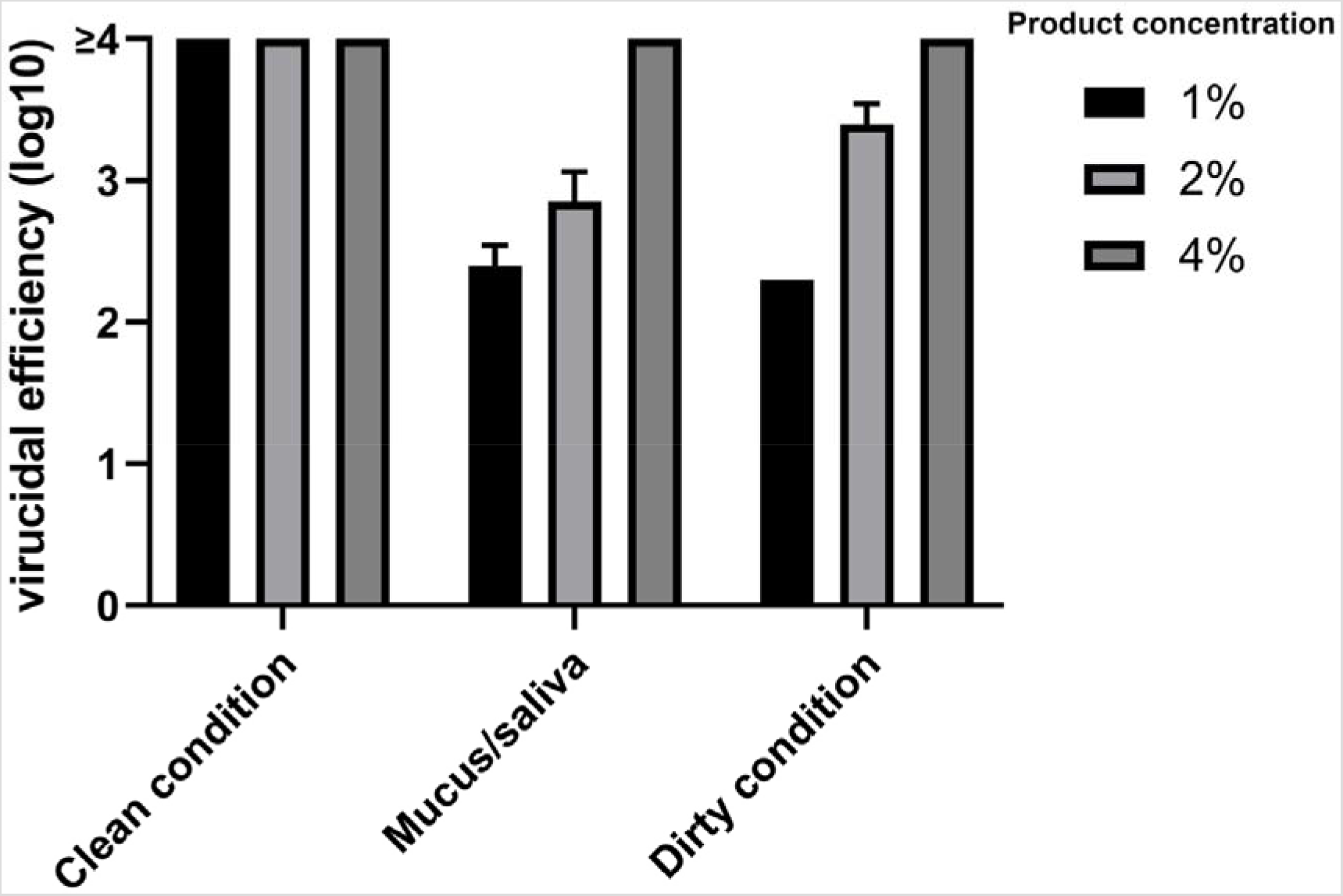
Virucidal activity of a commercial surface disinfectant. The purified virus was mixed with the interfering substance (“clean” condition: BSA 0.3 g/l, artificial saliva + epithelium mucus, “dirty” condition: BSA 3g/l + 3 ml/l of sheep erythrocytes). A volume of 50 µl of the mix was deposited on a previously functionalized stainless-steel disk (see previously). The inoculum was left to dry before 0.1 ml of the disinfectant (1, 2 and 4% concentrations) was deposited on the disk. The remaining infectious viruses were quantified by the Spearman and Karber method. The quantification limit was determined taking into account the cytotoxicity of the product (1.5 log10 TCID50).

## Discussion

The role of contaminated surfaces as a potential significant vector for hospital-acquired viral infections has gained a whole new community-based dimension in the current COVID-19 pandemic context. In that regard, the results presented in this study demonstrate the stability of SARS-CoV-2 on smooth surfaces, with infectious virus being detected up to 48 h and 72 h post application at 25 °C and 7 °C respectively. These observations confirm recent data that SARS-CoV-2 remains infectious for several days on stainless steel surface (10, 11). Our data suggest a higher stability of both HCoV-229E and FCoV at both temperatures compared to SARS-CoV-2, with infectious particles being detected up to 72 h post application as described previously (14). As previously described for coronavirus and other respiratory viruses, SARS-CoV-2 showed greatest surface-stability at low temperature (11, 12, 14). Numerous studies have described the role of body secretions in the observed increased stability of a large number of viral strains on environmental surfaces (15–17), as demonstrated for influenza virus in contact with respiratory mucus (18). Interestingly, the mucus/saliva mixture did not demonstrate a beneficial effect on surface survival for all tested viruses and a negative effect in SARS-CoV-2 seemed to correlate with the incubation temperature. At moderate temperatures, the mucus/saliva mixture had a strong impact on the viability of coronaviruses and quantities of virus close to the limit of detection (LOD) were detected up to 30 h. We hypothesize that the increase in temperature from 7 to 25 °C may activate the catalytic site of antimicrobial proteins present in nasal mucus. In this sense, the most abundant airway antimicrobial factors are lysozyme (31), lactoferrin (32), and secretory leukoproteinase inhibitor (33). Lactoferrin is a multifunctional glycoprotein with a broad spectrum of antiviral activity against a wide range of viruses *in vitro* including enveloped viruses (34), while the antiviral spectrum of lysozyme is considerably more modest (31). Several other known antimicrobial proteins and peptides including statherin and secretory phospholipase A2 have been identified in nasal secretions and likely contribute to their antimicrobial properties (34). As observed for rhinoviruses, nasal secretions and saliva seem to reduce the ability of coronaviruses to survive on environmental surfaces at moderate temperatures (20). However, we show that a large quantity of infectious SARSCoV-2 were still detectable on surfaces after 24 h post application at 25 °C in the presence of artificial body fluid secretions from coughing or sneezing. Laboratory simulations have demonstrated that hand-to-hand and fomite-to-hand contact are viable modes of transmission for influenza and other respiratory viruses depending how long the viral particles remain viable on hands and fomites (9, 24). As compared to influenza virus stability, we can hypothesize that SARS-CoV-2 remains stable on fomite as long as sufficient to contaminate hands and the risk of contaminating surfaces with SARSCoV-2 should be emphasized in guidelines for hand and surface disinfection practices.

With regards to disinfection, antiseptic hand hygiene including use of alcohol-based hand sanitizer is one of the most important measures in preventing outbreak-associated viral infections. We have shown that among the three commercially available alcohol hand sanitizers tested in our study, only one, corresponding to the ethanol-based WHO formulation, was associated with ≥4 log10 reduction after 30 s contact with SARS-CoV-2, HCoV-229E and FCoV, while the others reached compliance with EN14476 only after 60 s. This result confirms data from Siddharta et al. that ethanol-based WHO formulations efficiently inactivate re-emerging viral pathogens such as SARS-CoV, and MERS-CoV (35). However, the efficacy of such products is highly dependent on the technique of application, including the use of 2.4 to 3 ml applied to the palm and rubbed all over the surfaces of both hands until they are dry, corresponding to an acceptable contact time of 25 to 30 s (36). As the virucidal efficiency of alcohol-based hand sanitizers is determined according to EN14476 protocol, a contact time of 30 s should be considered only the reference to obtain compliance according to that standard. Noteworthy, we are currently performing a comparative study involving murine hepatitis virus (betacoronavirus), MVA virus, Type 5 adenovirus and murine norovirus to define the closest surrogate for SARS-CoV-2 to enable virucidal experiments with a contact time of 30 s on a large panel of alcohol-based hand sanitizers.

The mucus/saliva mixture used in this study showed a strong impact on virucidal efficiency when disinfectant was applied onto a dried inoculum-contaminated surface; in contrast however, it had no impact on virucidal efficiency tested using the suspension protocol with no significant difference shown between standardized clean medical condition and mucus/saliva mixture. We hypothesize that the dilution factor of the mucus/saliva mixture used in the modified suspension protocol at 97%, regularly used to evaluate ready-to-use hand rub products, may explain this difference. However, mucus/saliva interference had a similar impact on virucidal activity on surfaces as observed with the standardized medical dirty condition.

In summary, the democratization of the use of alcohol-based hand sanitizers from healthcare facilities to the general public as a barrier measure to prevent outbreak-associated respiratory viral infections emphasizes the importance of assessing the potential impact of interfering substances rich in protein or other biological organic matter, which are usually present in soiled hands and might significantly impact virucidal efficiency. Our results advocate for the development of standardized phase 2/step 2 testing protocols according to simulated practical conditions reflecting new “real life” needs, notably considering the impact of omnipresent secretions from coughing or sneezing on the evaluation of the virucidal activity of hand sanitizers.

## Data Availability

The data associated with this manuscript are available on request.

## Abbreviations

RH: relative humidity

## Acknowledgements

LZ is the holder of a CIFRE PhD thesis grant from the Agence Nationale de Recherche Technologique (ANRT). This study was funded by INSERM REACTing (REsearch & ACtion emergING infectious diseases), CNRS, and Mérieux research grants. The sponsors had no role in study design, collection, analysis and interpretation of data, manuscript writing, or in the decision to submit the article for publication.

